# Trends and Determinants of HIV Testing Uptake among Men Aged 15–59 Years in Zambia: A Multilevel Analysis of the Zambia Demographic and Health Surveys, 2007–2024

**DOI:** 10.64898/2026.02.05.26345700

**Authors:** Samson Shumba, Mwaka Hachisaala, Masha Maguswi, Womba Samudimu

## Abstract

HIV testing remains the primary entry point to HIV prevention, treatment, and care. Although Zambia has made substantial progress in its HIV response, men remain less consistently reached by HIV testing services. This study assessed trends and determinants of HIV testing uptake among men aged 15–59 years in Zambia using repeated nationally representative survey data.

We pooled men’s data from the Zambia Demographic and Health Surveys (ZDHS) conducted in 2007, 2013/14, 2018, and 2024. The outcome was HIV testing uptake, defined as ever tested for HIV and received results (DHS variable mv781). Analyses accounted for the complex survey design using sampling weights in Stata 14.2. Trends were examined using weighted proportions and design-adjusted chi-square tests. Determinants were assessed using multilevel logistic regression with men nested within clusters, reporting adjusted odds ratios (AORs) and 95% confidence intervals (CIs).

HIV testing uptake increased markedly from 20.7% (2007) to 62.9% (2013/14) and peaked at 77.4% (2018), with a modest decline in 2024 (73.5%); differences across survey years were statistically significant (p<0.05). In the fully adjusted model, survey year remained a strong predictor of testing compared with 2007 (2013/14 AOR 6.91, 95% CI 5.62–8.49; 2018 AOR 13.85, 95% CI 11.21–17.12; 2024 AOR 7.24, 95% CI 5.86–8.95). Older age was associated with higher odds of testing (25–34 AOR 3.51; 35–49 AOR 3.08; 50–59 AOR 1.65 vs 15–24). Rural residence was associated with lower testing (AOR 0.82, 95% CI 0.72–0.93). Higher education showed a strong gradient (primary AOR 1.55; secondary/higher AOR 4.19 vs none). Married men (AOR 4.33, 95% CI 3.56–5.27) and employed men (AOR 1.32, 95% CI 1.17–1.49) had higher odds of testing. Significant regional differences persisted after adjustment.

HIV testing uptake among men in Zambia rose substantially from 2007 to 2018 and remained high in 2024, though gaps persisted among younger men, rural residents, and selected provinces. Targeted, male-friendly strategies especially for younger and rural men and geographically tailored programming are needed to sustain gains and reduce inequities in HIV testing.

## Background

Human Immunodeficiency Virus (HIV) remains one of the most significant global public health challenges. As of the end of 2024, an estimated 40.8 million people were living with HIV worldwide, with approximately 1.3 million new infections and about 630,000 AIDS-related deaths reported in the same year [1,2]. Sub-Saharan Africa continues to shoulder a disproportionate share of the epidemic, accounting for the majority of people living with HIV globally [2]. In Zambia, nationally representative survey data indicate that HIV prevalence among adults aged 15–49 years is 8.7% overall, with markedly higher prevalence among women (11.1%) compared with men (5.9%) [3].

Globally, substantial progress has been made in expanding HIV testing and treatment coverage. As of 2024, an estimated 87% of people living with HIV were aware of their HIV status, and approximately 77% of those diagnosed were receiving antiretroviral therapy (ART), reflecting continued progress toward global HIV control targets [2]. In Zambia, the national HIV response has achieved notable success, with recent programmatic data indicating that the country has surpassed the UNAIDS 95–95–95 targets, attaining cascade achievements of 98–98–97 across diagnosis, treatment, and viral suppression [1].

Despite these gains, men remain a persistently underserved population within HIV programmes, particularly with respect to HIV testing uptake [4–6]. Evidence from sub-Saharan Africa consistently demonstrates lower HIV testing coverage among men compared with women, driven by a combination of gender norms, reduced health-care engagement, and structural barriers to service access [4–8]. For example, in Mozambique, 38.3% of men have ever tested for HIV compared with 47.6% of women [9,10]. Across the region, male HIV testing uptake varies widely, ranging from as low as 10.2% in Guinea and 18.7% in Nigeria to substantially higher levels observed in Cameroon (58%) and Rwanda (65%), highlighting persistent cross-country and gender-based inequities in HIV service utilization [11–13].

HIV testing is a critical step in the prevention of HIV transmission, treatment, care, and other supportive services [14]. Early detection allows for timely initiation of ART, which substantially reduces morbidity, mortality, and onward transmission. Furthermore, testing enables HIV-negative individuals to access prevention resources. However, men remain a hard-to-reach population with respect to HIV testing uptake, posing a continued challenge to achieving equitable HIV service coverage [4].

Existing evidence indicates that men are less likely than women to seek HIV testing services, resulting in delayed diagnosis and weaker engagement across the HIV care cascade [4,15,16]. Studies from sub-Saharan Africa show that HIV testing uptake among men is strongly influenced by individual and socio-economic factors, including HIV/AIDS knowledge, age, education, employment, and household wealth, with higher awareness and socio-economic status associated with greater testing uptake [17–19]. Country-specific evidence further highlights the role of marital status, religion, male circumcision, media exposure, and region of residence in shaping men’s HIV testing behaviour, while engagement in HIV-related risk behaviours has been linked to lower recent testing uptake [15,16,20–22]. In addition, social norms, stigma, and dominant masculinity ideals remain important barriers to HIV testing among men, with fear of knowing one’s HIV status frequently discouraging service utilization [4,9,23]. Nonetheless, facilitating factors such as family responsibilities, including fatherhood, have been shown to motivate HIV testing among men in Zambia and other settings [24].

Although several studies have examined determinants of HIV testing among men in sub-Saharan Africa, fewer have assessed how these patterns have evolved over time using repeated nationally representative surveys. In Zambia, existing evidence has largely focused on women or the general population, with limited analysis of long-term trends and determinants of HIV testing uptake specifically among men. Given the critical importance of reaching men to sustain Zambia’s progress toward epidemic control, this study aims to examine trends and determinants of HIV testing uptake among men aged 15–59 years in Zambia using data from the Zambia Demographic and Health Surveys conducted between 2007 and 2024.

## Methodology

### Study Population and Study Setting

This study used data from the Zambia Demographic and Health Surveys (ZDHS) conducted in 2007, 2013/14, 2018, and 2024. The ZDHS are nationally representative household surveys designed to generate reliable estimates of key demographic, health, and HIV-related indicators at both national and subnational levels. Zambia is administratively divided into ten provinces, each comprising urban and rural areas, which served as the primary domains of analysis. The study population consisted of men aged 15–59 years who were interviewed in the men’s individual survey across the four DHS rounds. Eligible respondents were those with complete information on HIV testing history and relevant socio-demographic and behavioural characteristics.

The ZDHS employed a two-stage stratified sampling design. In the first stage, enumeration areas (EAs) were selected as primary sampling units from the 2010 Census of Population and Housing. In the second stage, households were systematically selected within each EA, and all eligible men aged 15–59 years in the selected households were interviewed. Sampling weights provided by the DHS program were applied in all analyses to account for differential probabilities of selection and non-response, thereby ensuring national representativeness. The surveys were implemented by the Zambia Statistics Agency in collaboration with the Ministry of Health Zambia and the University Teaching Hospital Virology Laboratory, with technical assistance from ICF through the Demographic and Health Surveys Program.

### Study Variables and Their Definitions

The outcome variable for this study was HIV testing uptake, defined as whether a respondent had ever been tested for HIV and received the test results, measured using the DHS variable mv781 (*“Ever tested for HIV and received results”*). Explanatory variables comprised both individual-and community-level factors. Individual-level variables included age group (mv013), highest educational attainment (mv106), household wealth index (mv190), media exposure (derived from mv157, mv158, and mv159), marital status (mv501), employment status (mv731), HIV knowledge score (constructed from mv754a–mv754j), awareness of sexually transmitted infections (mv763a), history of sexually transmitted infections (mv763b–mv763d), and adolescent fatherhood status (derived from mv201 and age at first birth). Community-level variables were constructed at the cluster level (mv021) and included aggregated measures of poverty (wealth index), educational attainment, and employment status. These variables were selected to capture both individual characteristics and broader contextual influences on HIV testing uptake among men in Zambia.

**Table 1:** Definition of Dependent and Independent Variables.

| Variable | Type | Coding / Categories | Level | Description |
| --- | --- | --- | --- | --- |
| HIV Test Uptake | Dependent | 0 = Not tested; 1 = Tested & received results | Individual | Indicates whether respondent has ever tested for HIV and received results |
| Age Group | Independent | 1 = 15–24; 2 = 25–34; 3 = 35–49; 4 = 50–59 | Individual | Respondent's age categorized |
| Education | Independent | 1 = None; 2 = Primary; 3 = Secondary/Higher | Individual | Highest education level attained |
| Wealth Index | Independent | 1 = Poor; 2 = Middle; 3 = Rich | Individual | Socioeconomic status based on household wealth |
| Media Exposure | Independent | 0 = No; 1 = Yes | Individual | Exposure to radio, newspaper, or TV |
| Marital Status | Independent | 0 = Not married; 1 = Currently married/in union | Individual | Current marital status |
| Employment Status | Independent | 0 = Not employed; 1 = Employed | Individual | Current employment status |
| HIV Knowledge Score | Independent | 0–4 | Individual | Score based on correct responses to HIV knowledge questions |
| STI Awareness | Independent | 0 = No; 1 = Yes | Individual | Knowledge of sexually transmitted infections |
| STI History | Independent | 0 = No STI/symptoms; 1 = Had STI/symptoms | Individual | Self-reported STI history |
| Adolescent Mother | Independent | 0 = Age 20–49; 1 = Age 15–19 | Individual | Indicator for adolescent age (if applicable) |
| Community Poverty | Independent | 0 = Low; 1 = Medium; 2 = High | Community | Cluster-level mean proportion of households in poverty |
| Community Education | Independent | 0 = Low; 1 = Medium; 2 = High | Community | Cluster-level mean proportion of respondents with low education |
| Community Employment Status | Independent | 0 = Low; 1 = Medium; 2 = High | Community | Cluster-level mean proportion of respondents not employed |

### Data Analysis

Data from the Zambia Demographic and Health Surveys (ZDHS) conducted in 2007, 2013/14, 2018, and 2024 were pooled and analysed to examine temporal trends and determinants of HIV testing uptake among men aged 15–59 years. All analyses were performed using Stata version 14.2. Sampling weights were applied to account for the complex survey design and to ensure nationally representative estimates. Descriptive statistics were first generated to summarise the distribution of socio-demographic characteristics and HIV testing uptake across survey years. Bivariate associations between HIV testing uptake and selected socio-demographic factors were assessed using chi-square tests of independence, with statistical significance evaluated at the 5% level (p < 0.05).

To identify determinants of HIV testing uptake, multilevel logistic regression models were fitted to account for the hierarchical structure of the data, with individuals nested within clusters and regions. The outcome variable was HIV testing uptake (ever tested for HIV and received results versus never tested). Individual-level explanatory variables included age group (15–24, 25–34, 35–49, 50–59 years), educational attainment (none, primary, secondary or higher), marital status (currently married/in union versus not married), employment status (employed versus not employed), media exposure (exposed versus not exposed), HIV knowledge score (0–4), and awareness of sexually transmitted infections (STIs; aware versus not aware). Community-level variables comprised aggregated cluster-level measures of poverty, education, and employment status.

Three multilevel models were estimated sequentially. Model 1 (the null model) assessed the extent of clustering in HIV testing uptake. Model 2 included individual-level variables only, while Model 3 (the full model) incorporated both individual- and community-level variables. Results are presented as adjusted odds ratios (ORs) with corresponding 95% confidence intervals (CIs). Model fit was evaluated using the Akaike Information Criterion (AIC), Bayesian Information Criterion (BIC), and the intra-class correlation coefficient (ICC), which quantified the proportion of variance attributable to cluster-level effects. Graphical analyses, including bar charts, were used to illustrate patterns of HIV testing uptake across provinces and survey years. All analyses accounted for survey weights and design effects to produce valid population-level estimates.

### Ethical Clearance

This study utilized secondary data obtained from the Zambia Demographic and Health Surveys (ZDHS) conducted in 2007, 2013/14, 2018, and 2024. The datasets are publicly available and were accessed for research purposes on 15 January 2026 following formal request and approval from the DHS Program (www.dhsprogram.com). The surveys were implemented by the Zambia Statistics Agency (ZamStats) in collaboration with the Ministry of Health (MOH) and the University Teaching Hospital (UTH) Virology Laboratory, with technical assistance from ICF International. All ZDHS protocols were reviewed and approved by the Tropical Diseases Research Centre (TDRC) Ethics Review Committee in Zambia and the ICF Institutional Review Board (IRB) in the United States. Written informed consent was obtained from all participants prior to survey implementation.

## Results

### Trends in HIV Test Uptake among Men in Zambia: 2007–2024

The analysis of HIV testing uptake across the Zambia Demographic and Health Surveys (ZDHS) for 2007, 2013–14, 2018, and 2024 shows a substantial increase in the proportion of respondents who tested for HIV and received their results. In 2007, only 20.7% of individuals reported having been tested and received their test results. This proportion rose sharply to 62.9% in the 2013–14 survey. By 2018, HIV testing uptake further increased to 77.4%. Although there was a slight decline in 2024, a high proportion 73.5% of respondents still reported having tested and received their results. A chi-square test of association, adjusted for the complex survey design, indicated that the differences in HIV testing uptake across the survey years were statistically significant (see Fig 1 below).

**Fig 1:**
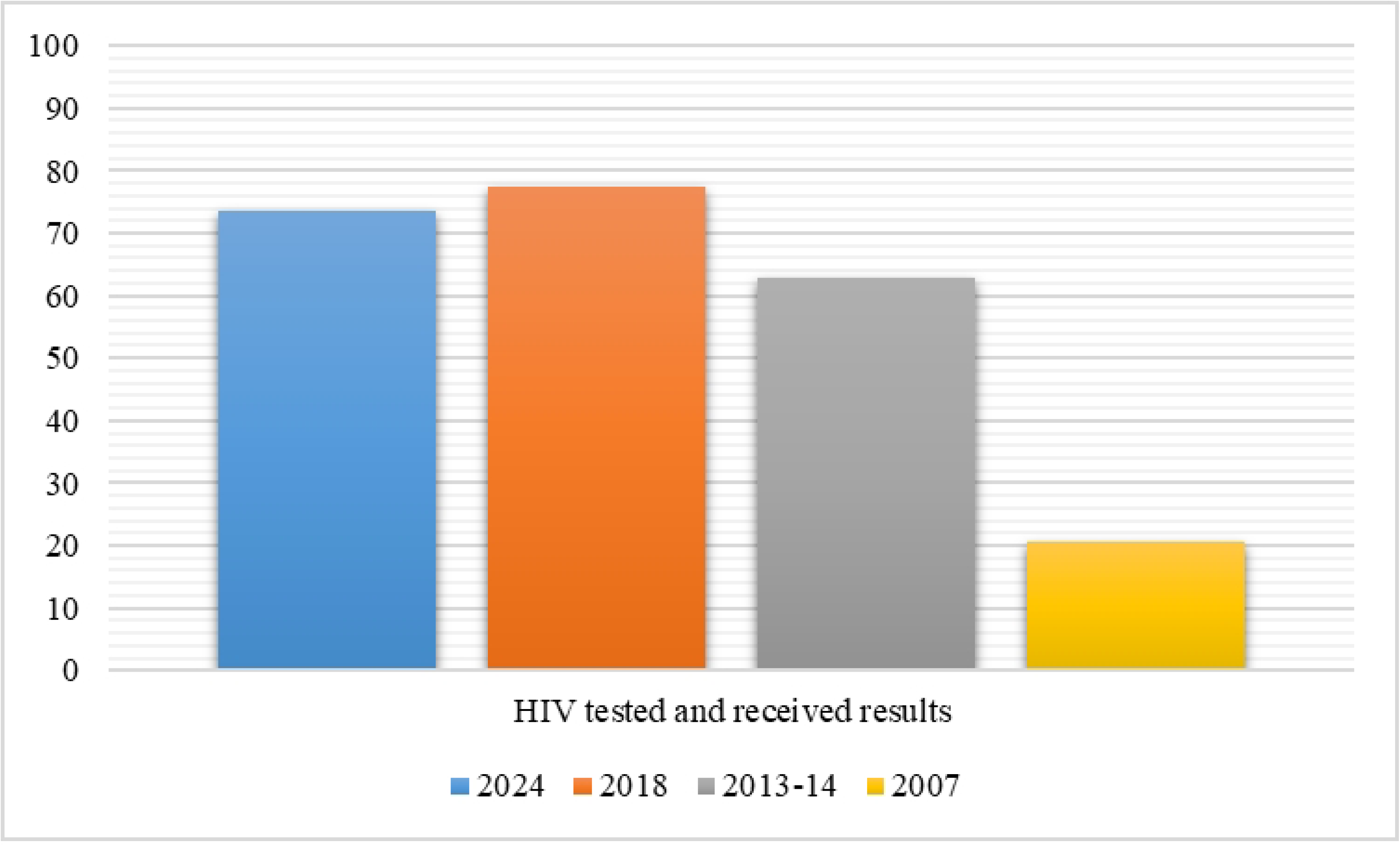
Percentage of Men Who Have Tested and Received HIV Results across Survey Years (ZDHS 2007, 2013–14, 2018 and 2024)

### HIV Testing Uptake among Men by Region across Survey Years (ZDHS 2007–2024)

The analysis of HIV testing uptake across regions in Zambia shows clear improvements in the proportion of individuals who reported having been tested and received their HIV results between 2007 and 2024. In 2007, testing uptake was generally low across all regions, with the highest proportions observed in Lusaka (26.46%), Southern (24.26%), Central (24.06%), and Western (21.92%). The lowest levels were recorded in Northern (13.70%), Luapula (15.38%), and Eastern (16.99%) provinces. By the 2013–14 survey, there were marked improvements across nearly all regions. HIV testing uptake increased to between 53% and 76%. Western Province recorded the highest uptake at 76.05%, followed by Southern (67.65%), Eastern (67.12%), and Luapula (64.68%). The lowest proportions were seen in Northern (53.39%) and Central (54.55%).

In 2018, HIV testing uptake continued to rise, reaching the highest levels observed since 2007. Southern Province recorded the highest uptake (84.61%), followed by Lusaka (81.82%), Western (80.08%), and Eastern (79.11%). Muchinga (65.73%), Luapula (69.04%), and Copperbelt (74.02%) registered comparatively lower but still substantial improvements. In 2024, HIV testing uptake remained high across most regions, although some provinces showed slight declines compared to 2018. The highest proportions were recorded in Lusaka (80.40%), Southern (77.67%), Central (76.62%), and Western (77.35%). Lower levels were observed in North-Western (65.14%), Muchinga (66.43%), and Northern (67.51%), see Fig 2 below.

**Fig 2:**
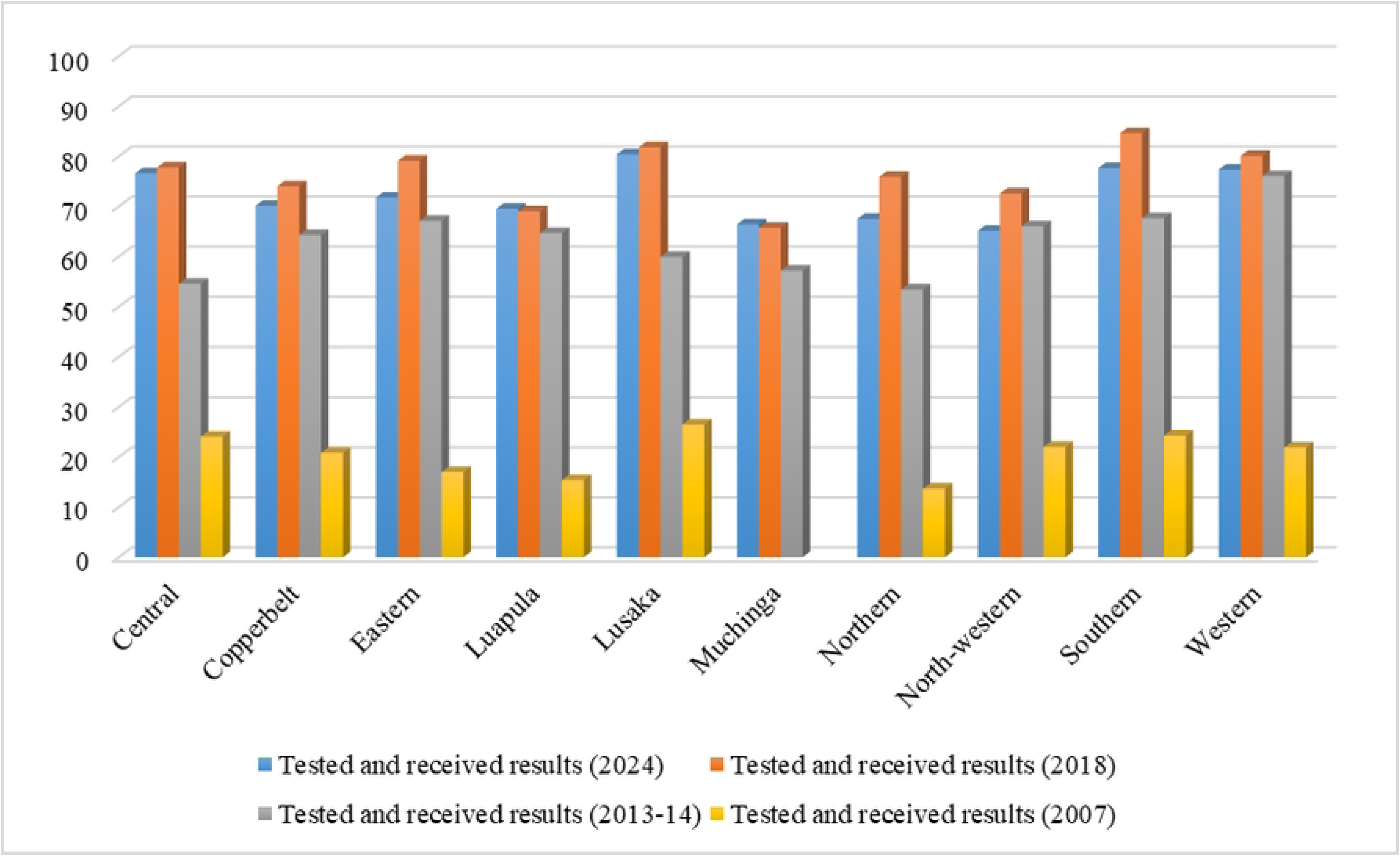
HIV Test Uptake among Men by Province cross Survey Years (ZDHS 2007–2024)

### HIV Test Uptake among Men by Selected Background Characteristics across ZDHS 2007, 2013–14, 2018 and 2024

A comparison of HIV testing uptake among men across the Zambia Demographic and Health Surveys (ZDHS) from 2007, 2013–14, 2018, and 2024 reveals substantial improvements over time, with statistically significant differences observed across survey years (p < 0.05). In 2007, HIV testing uptake was low across nearly all socio-demographic groups. Uptake increased sharply in 2013–14, continued rising in 2018, and remained relatively high in 2024 despite slight declines in some subgroups.

Across all years, age showed consistent increases in uptake. In 2007, fewer than 26% of men aged 25–49 had ever tested for HIV, and uptake among younger men (15–24) was especially low (14.7%). By 2013–14 and 2018, testing increased across all age groups, reaching its highest levels in 2018. In 2024, uptake remained high among older men (89–93%) but declined notably among younger men (47.8%), suggesting that testing gains were not evenly sustained. Residence showed similar upward progression. Urban and rural men both exhibited low uptake in 2007 (24.5% and 18.0%, respectively). Uptake rose markedly through 2013–14 and peaked in 2018 (80.6% urban; 75.1% rural). In 2024, both groups experienced slight decreases (76.2% urban; 71.1% rural), although levels remained far higher than in earlier years.

Across survey years, education demonstrated a strong upward trend. In 2007, uptake ranged from 13.1% among men with no education to 27.1% among those with secondary or higher education. Significant increases were observed in 2013–14 and 2018, when uptake exceeded 80% among the most educated. In 2024, testing remained highest among men with secondary or higher education (79.3%) but was lower than 2018 levels. Similarly, wealth index displayed year-on-year improvements. In 2007, only 15.1% of the poorest men and 24.9% of the richest men had tested. Testing increased substantially in 2013–14 and 2018 across all wealth groups. Although uptake declined slightly in 2024, the rich (77.2%) and poor (70.5%) remained far above 2007 levels.

Across survey years, marital status showed one of the strongest comparative trends. Married men consistently exhibited higher uptake than unmarried men. In 2007, less than one-third of married men had tested. By 2018, uptake exceeded 90%, and in 2024 this pattern continued (92.8% among married men vs. 53.4% among unmarried men). Media exposure, employment, HIV knowledge, and STI awareness all showed similar upward trajectories between 2007 and 2018, followed by slight reductions in 2024. However, uptake in 2024 remained significantly higher than in the earliest years. Knowledge-related indicators, particularly HIV knowledge score, demonstrated the most consistent dose–response trend across survey years, with higher knowledge associated with significantly higher testing in all rounds.

**Table 2:** HIV Test Uptake among Men by Selected Background Characteristics across ZDHS 2007, 2013–14, and 2018.

| Variables | HIV test Uptake among Men (Overall) |  | HIV test Uptake among Men (2024) |  | HIV test Uptake among Men (2018) |  | HIV test Uptake among Men (2013-14) |  | HIV test Uptake among Men (2007) |  |
| --- | --- | --- | --- | --- | --- | --- | --- | --- | --- | --- |
|  | Not tested | Tested and received results | Not tested | Tested and received results | Not tested | Tested and received results | Not tested | Tested and received results | Not tested | Tested and received results |
| **Age** | **<0.0001** |  | **<0.0001** |  | **0.0001** |  | **0.0001** |  | **<0.0001** |  |
| 15 to 24 | 8774 (53.69) | 7567 (46.31) | 2558 (52.24) | 2338 (47.76) | 1780 (39.47) | 2730 (60.53) | 2828 (56.00) | 2223 (44.00) | 1608 (85.31) | 277 (14.69) |
| 25 to 34 | 2462 (23.10) | 8197 (76.90) | 296 (9.97) | 2673 (90.02) | 303 (10.47) | 2591 (89.53) | 782 (23.40) | 2562 (76.60) | 1080 (74.41) | 372 (25.59) |
| 35 to 49 | 2329 (21.61) | 8449 (78.39) | 215 (7.04) | 2836 (92.97) | 301 (10.07) | 2686 (89.93) | 857 (24.48) | 2645 (75.52) | 957 (77.20) | 282 (22.80) |
| 50 to 59 | 911 (26.68) | 2502 (73.32) | 111 (10.23) | 974 (89.77) | 161 (18.14) | 725 (81.86) | 341 (32.54) | 706 (67.46) | 298 (75.34) | 98 (24.66) |
| **Residence** | **<0.0001** |  | **<0.0001** |  | **<0.0001** |  | **0.0166** |  | **0.0166** |  |
| Urban | 5866 (32.07) | 12427 (67.93) | 1328 (23.78) | 4256 (76.22) | 949 (19.43) | 3933 (80.57) | 2042 (35.36) | 3734 (64.64) | 1548 (75.48) | 503 (24.52) |
| Rural | 8610 (37.60) | 14290 (62.40) | 1852 (28.86) | 4565 (71.14) | 1595 (24.95) | 4798 (75.05) | 2767 (38.60) | 4401 (61.40) | 2396 (82.00) | 526 (18.00) |
| **Education** | **<0.0001** |  | **<0.0001** |  | **<0.0001** |  | **<0.0001** |  | **<0.0001** |  |
| None | 751 (45.11) | 914 (54.89) | 204 (39.41) | 314 (60.59) | 140 (30.83) | 313 (69.17) | 208 (44.76) | 257 (55.24) | 199 (86.88) | 30 (13.12) |
| Primary | 7113 (43.03) | 9418 (56.97) | 1535 (33.91) | 2992 (66.09) | 1341 (30.31) | 3085 (69.69) | 2262 (42.98) | 3002 (57.02) | 1974 (85.32) | 340 (14.68) |
| Sec/Higher | 6606 (28.74) | 16381 (71.26) | 1440 (20.70) | 5515 (79.30) | 1063 (16.62) | 5334 (83.38) | 2332 (32.36) | 4873 (67.64) | 1771 (72.88) | 659 (27.12) |
| **Region** | **<0.0001** |  | **<0.0001** |  | **<0.0001** |  | **<0.0001** |  | **<0.0001** |  |
| Central | 1454 (35.32) | 2663 (64.68) | 343 (23.38) | 1123 (76.62) | 229 (22.21) | 801 (77.79) | 521 (45.45) | 625 (54.55) | 362 (75.94) | 115 (24.06) |
| Copperbelt | 2378 (37.42) | 3977 (62.58) | 498 (29.85) | 1170 (70.15) | 435 (25.98) | 1238 (74.02) | 773 (35.70) | 1392 (64.30) | 673 (79.17) | 177 (20.83) |
| Eastern | 1884 (34.95) | 3507 (65.05) | 436 (28.22) | 1110 (71.78) | 314 (20.89) | 1188 (79.11) | 532 (32.88) | 1086 (67.12) | 602 (83.01) | 123 (16.99) |
| Luapula | 1097 (38.39) | 1760 (61.61) | 262 (30.43) | 600 (69.57) | 250 (30.96) | 557 (69.04) | 302 (35.32) | 552 (64.68) | 283 (84.62) | 51 (15.38) |
| Lusaka | 2484 (32.24) | 5220 (67.76) | 402 (19.60) | 1647 (80.40) | 395 (18.18) | 1776 (81.82) | 1048 (40.08) | 1566 (59.92) | 640 (73.54) | 230 (26.46) |
| Muchinga | 1284 (50.35) | 1266 (49.65) | 186 (33.57) | 367 (66.43) | 216 (34.27) | 415 (65.73) | 292 (42.80) | 390 (57.20) |  |  |
| Northern | 1152 (38.18) | 1865 (61.82) | 291 (32.49) | 605 (67.51) | 220 (24.08) | 694 (75.92) | 446 (46.61) | 511 (53.39) | 590 (86.30) | 94 (13.70) |
| North-western | 992 (41.65) | 1390 (58.35) | 291 (32.49) | 511 (65.14) | 153 (27.43) | 405 (72.57) | 180 (33.98) | 351 (66.02) | 194 (77.98) | 55 (22.02) |
| Southern | 1317 (27.08) | 3546 (72.92) | 324 (22.33) | 1126 (77.67) | 215 (15.39) | 1182 (84.61) | 563 (32.35) | 1178 (67.65) | 385 (75.74) | 123 (24.26) |
| Western | 435 (22.25) | 1522 (77.75) | 165 (22.65) | 563 (77.35) | 118 (19.92) | 476 (80.08) | 152 (23.95) | 483 (76.05) | 215 (78.08) | 60 (21.92) |
| **Wealth Index** | **<0.0001** |  | **<0.0001** |  | **0<0.0001** |  | **0.0217** |  | **0.0217** |  |
| Poor | 5530 (38.27) | 8922 (61.73) | 1302 (29.51) | 3110 (70.49) | 1064 (27.28) | 2837 (72.72) | 1700 (38.50) | 2715 (61.50) | 1464 (84.89) | 261 (15.11) |
| Middle | 2857 (35.26) | 5247 (64.74) | 699 (28.74) | 1733 (71.26) | 468 (20.51) | 1812 (79.49) | 967 (38.96) | 1515 (61.04) | 724 (79.47) | 187 (20.53) |
| Rich | 6089 (32.67) | 12547 (67.33) | 1179 (22.85) | 3979 (77.15) | 1012 (19.87) | 4083 (80.13) | 2142 (35.43) | 3904 (64.57) | 1756 (75.14) | 581 (24.86) |
| **Media Exposure** | **0.0329** |  | **0.0027** |  | **<0.0001** |  | **0.4178** |  | **0.4178** |  |
| No exposure | 10102 (35.61) | 18270 (64.39) | 2375 (27.36) | 6305 (72.64) | 1859 (23.90) | 5919 (76.10) | 3112 (36.82) | 5340 (63.18) | 2756 (79.62) | 705 (20.38) |
| Exposed | 4374 (34.12) | 8446 (65.88) | 805 (24.23) | 2516 (75.77) | 685 (19.59) | 2812 (80.41) | 1697 (37.78) | 2795 (62.22) | 1188 (78.60) | 323 (21.40) |
| **Current Marital Status** | **<0.0001** |  | **<0.0001** |  | **<0.0001** |  | **<0.0001** |  | **<0.0001** |  |
| Not married | 9664 (50.32) | 9543 (49.68) | 2737 (46.56) | 3142 (53.44) | 1962 (37.00) | 3341 (63.00) | 3163 (54.20) | 2673 (45.80) | 1803 (82.30) | 388 (17.70) |
| Currently married | 4812 (21.89) | 17173 (78.11) | 443 (7.24) | 5680 (92.76) | 582 (9.74) | 5391 (90.26) | 1646 (23.16) | 5462 (76.84) | 2141 (76.96) | 641 (23.04) |
| **Employment Status** | **<0.0001** |  | **<0.0001** |  | **<0.0001** |  | **<0.0001** |  | **0.3115** |  |
| Not employed | 4613 (54.56) | 3842 (45.44) | 1405 (52.12) | 1291 (47.88) | 984 (42.06) | 1356 (57.94) | 1413 (57.04) | 1064 (42.96) | 810 (86.13) | 131 (13.87) |
| Employed | 898 (36.46) | 1565 (63.54) | 293 (31.46) | 637 (68.54) | 102 (22.92) | 341 (77.08) | 357 (39.05) | 557 (60.95) | 147 (83.05) | 30 (16.95) |
| **HIV knoweldge score** | **<0.0001** |  | **<0.0001** |  | **<0.0001** |  | **<0.0001** |  | **0.0004** |  |
| 0 | 916 912.31) | 6523 (87.69) | 681 (9.49) | 6501 (90.51) | 116 (92.43) | 9 (7.57) | 73 (90.81) | 7 (9.19) | 45 (89.39) | 5 (10.61) |
| 1 | 490 (67.60) | 235 (32.40) | 109 (72.79) | 41 (27.21) | 62 (50.04) | 62 (49.96) | 148 (58.14) | 107 (41.86) | 171 (86.89) | 26 (13.11) |
| 2 | 1183 (54.20) | 999 (45.80) | 279 (60.95) | 179 (39.05) | 146 (30.34) | 336 (69.66) | 380 (48.74) | 400 (51.26) | 377 (81.05) | 84 (18.29) |
| 3 | 3699 (47.08) | 4158 (52.92) | 733 (54.03) | 624 (45.97) | 634 (28.10) | 1623 (71.90) | 1144 (41.19) | 1634 (58.81) | 1187 (81.05) | 278 (18.95) |
| 4 | 8189 (35.62) | 14801 (64.38) | 1377 (48.25) | 1477 (51.75) | 1586 (19.14) | 6701 (80.86) | 3064 (33.85) | 5988 (66.15) | 2163 (77.29) | 635 (22.71) |
| **Aware of STIs** | **<0.0001** |  | **0.0009** |  | **<0.0001** |  | **0.0009** |  | **<0.0001** |  |
| No | 14008 (35.39) | 25570 (64.61) | 3038 (26.67) | 8352 (73.34) | 2498 (22.90) | 8410 (77.10) | 4679 (37.44) | 7819 (62.56) | 3793 (79.30) | 990 (20.70) |
| Yes | 418 (27.44) | 1104 (72.56) | 120 (20.78) | 458 (79.22) | 43 (11.84) | 322 (88.16) | 106 (27.16) | 285 (72.84) | 148 (79.33) | 38 (20.67) |
| **Community Poverty** | **0.7517** |  | **0.0003** |  | **<0.0001** |  | **0.47** |  |  |  |
| Low | 11198 (34.91) | 20877 (65.09) | 2024 (26.22) | 5696 (73.78) | 1387 (20.87) | 5258 (79.13) | 3053 (37.46) | 5097 (62.54) | 1782 (79.84) | 450 (20.16) |
| Medium | 2566 (35.92) | 4577 (64.08) | 407 (29.11) | 990 (70.89) | 453 (28.93) | 1114 (71.07) | 564 (37.59) | 936 (62.41) | 448 (84.16) | 84 (15.84) |
| High | 538 (36.16) | 950 (63.84) | 306 (33.31) | 612 (66.69) | 176 (32.66) | 362 (67.34) | 242 (41.73) | 337 (58.27) | 475 (86.89) | 72 (13.11) |
| **Community Education** | **0.3047** |  | **<0.0001** |  | **<0.0001** |  | **0.0349** |  | **<0.0001** |  |
| Low | 12329 (34.84) | 23056 (65.16) | 2170 (24.93) | 6532 (75.07) | 1695 (20.72) | 6486 (79.28) | 3181 (36.16) | 5615 (63.84) | 2180 (76.11) | 684 (23.89) |
| Medium | 1798 (36.79) | 3089 (63.21) | 580 (29.10) | 1414 (70.90) | 459 (26.14) | 1297 (73.86) | 928 (37.61) | 1539 (62.39) | 942 (82.17) | 204 (17.84) |
| High | 350 (37.97) | 571 (62.03) | 430 (32.94) | 875 (67.06) | 390 (29.14) | 949 (70.86) | 700 (41.66) | 981 (58.34) | 822 (85.45) | 140 (14.55) |
| **Community**<br>**Employment**<br>**Status** | **0.7428** |  | **<0.0001** |  | **0.9376** |  | **0.0349** |  | **<0.0001** |  |
| Low | 665 (33.66) | 1312 (66.34) | 2170 (24.93) | 6532 (75.07) | 152 (21.86) | 543 (78.14) | 698 (34.61) | 1318 (65.39) | 306 (81.28) | 70 (18.72) |
| Medium | 996 (34.95) | 1854 (65.05) | 580 (29.10) | 1414 (70.90) | 150 (22.80) | 507 (77.20) | 436 (40.44) | 642 (59.56) | 232 (80.10) | 58 (19.90) |
| High | 12768 (35.25) | 23452 (64.75) | 430 (32.94) | 875 (67.06) | 2156 (22.65) | 7365 (77.35) | 3385 (37.35) | 5679 (62.83) | 2932 (78.82) | 933 (21.18) |

### Multilevel Analysis of HIV Test Uptake among Men in Zambia

The multilevel logistic regression model (Model 3) examined factors associated with HIV testing uptake among men across the Zambia Demographic and Health Surveys (ZDHS) from 2007, 2013/14, and 2018. This model included both individual and community-level variables. Survey year remained a strong predictor, with individuals surveyed in 2013 (OR: 6.91, 95% CI: 5.62–8.49), 2018 (OR: 13.85, 95% CI: 11.21–17.12), and 2024 (OR: 7.24, 95% CI: 5.86–

8.95) having significantly higher odds of undergoing HIV testing compared to those surveyed in 2007, holding all other variables constant. Age was also significantly associated with HIV testing, where respondents aged 25–34 and 35–49 had more than three times higher odds of testing relative to those aged 15–24, while adults aged 50–59 also showed elevated odds (OR: 1.65, 95% CI: 1.22–2.22). Rural residence was associated with reduced likelihood of HIV testing (OR: 0.82, 95% CI: 0.72–0.93) compared to urban areas.

Education demonstrated a clear gradient effect after controlling for other covariates: individuals with primary education (OR: 1.55, 95% CI: 1.20–2.00) and those with secondary or higher education (OR: 4.19, 95% CI: 3.21–5.47) were significantly more likely to test for HIV than those with no formal education. Substantial regional variations persisted even after adjusting for all individual-level characteristics. Provinces such as Copperbelt, Muchinga, Northern, and Northwestern had significantly lower odds of HIV testing compared with Central Province, whereas Western Province exhibited higher odds (OR: 1.72, 95% CI: 1.37–2.15). Employment status was positively associated with HIV testing uptake (OR: 1.32, 95% CI: 1.17–1.49), while marital status emerged as one of the strongest predictors, with married individuals having more than four times higher odds of testing than those not married (OR: 4.33, 95% CI: 3.56–5.27), holding all other factors constant.

HIV knowledge scores displayed a distinct dose–response relationship, where increasing knowledge was associated with progressively higher odds of testing. In contrast, none of the community-level variables community poverty, education, or employment—showed significant associations in the fully adjusted model, suggesting that individual-level characteristics had greater explanatory power for HIV testing uptake. Model diagnostics demonstrated improved performance for Model III, which had the lowest AIC (11705.24) and BIC (11966.81) values. The intra-class correlation coefficient (ICC = 0.012, 95% CI: 0.004–0.032) indicated minimal residual clustering at the community level after accounting for both individual and contextual factors.

**Table 3:**
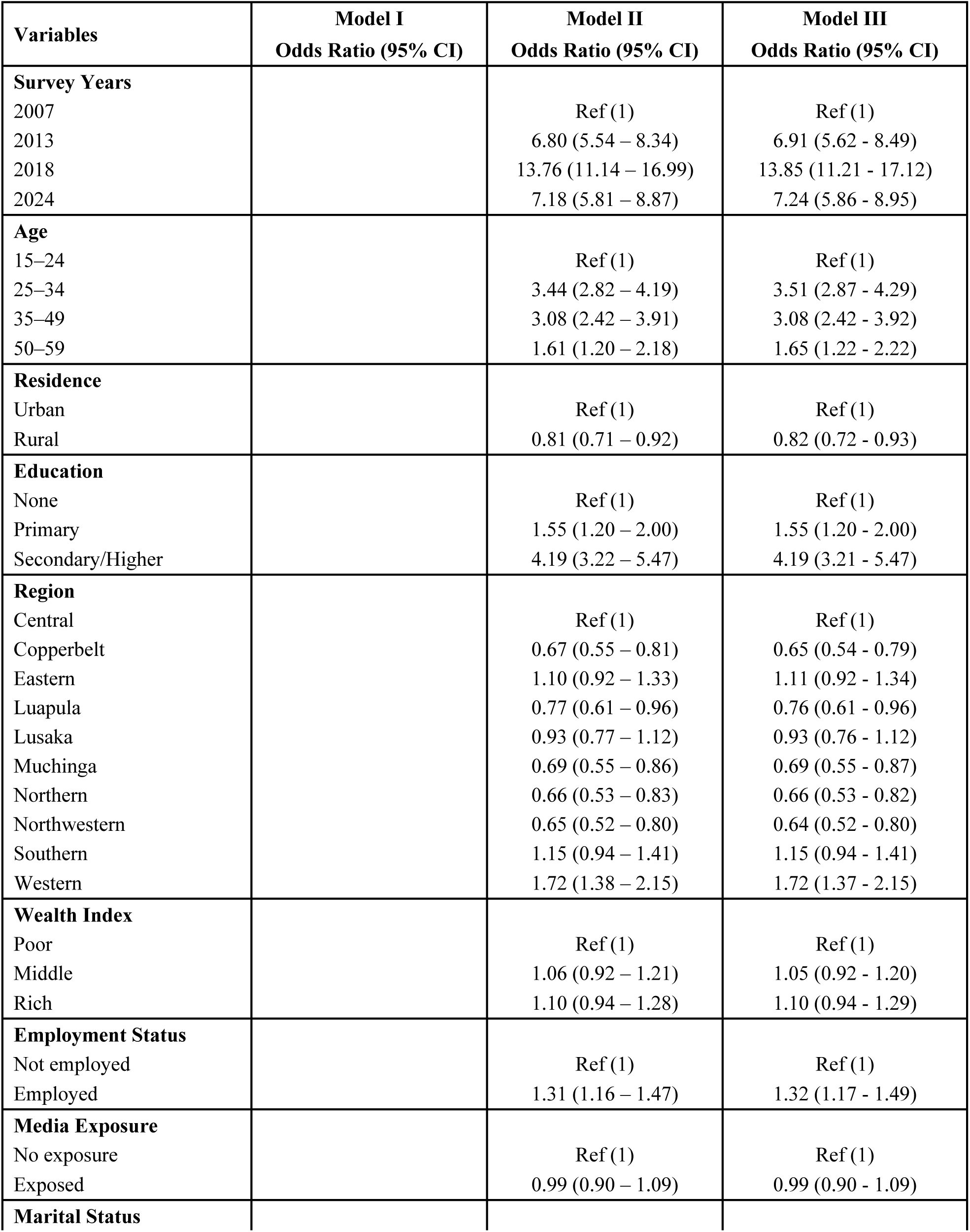
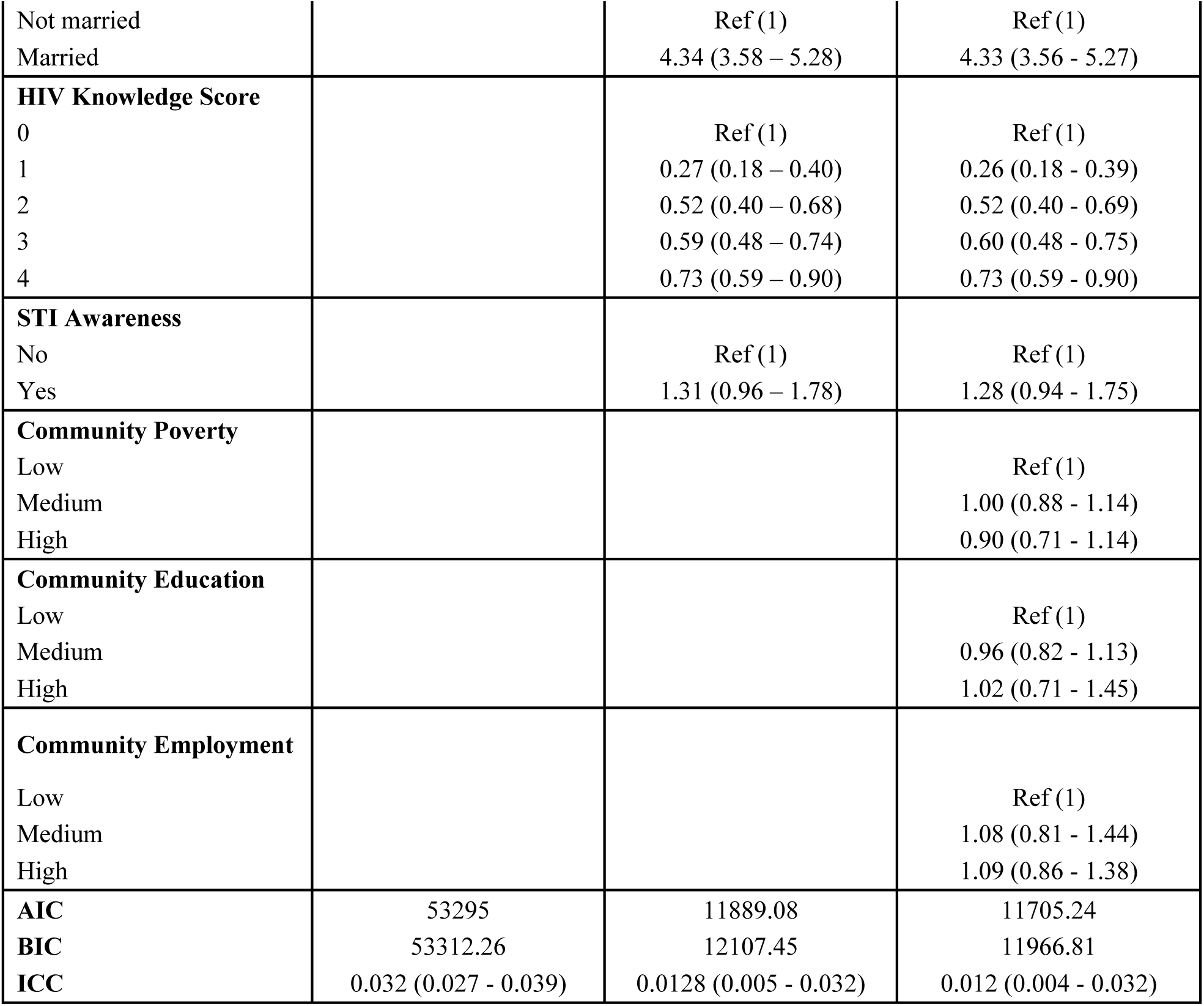
Multilevel Logistic Regression of HIV Test Uptake among Men in Zambia (ZDHS 2007, 2013/14, 2018 and 2024)

## Discussion

This study examined trends and determinants of HIV testing uptake among men aged 15–59 years in Zambia using nationally representative data from four rounds of the Zambia Demographic and Health Surveys (ZDHS) spanning 2007–2024. The findings demonstrate a substantial and statistically significant increase in HIV testing uptake over time, with testing prevalence rising from 20.7% in 2007 to 62.9% in 2013–14 and peaking at 77.4% in 2018. Although a modest decline was observed in 2024 (73.5%), testing uptake remained markedly higher than levels recorded in the pre-scale-up period. This pattern reflects sustained progress in male HIV testing in Zambia and is consistent with trends reported in other sub-Saharan African countries following large-scale investments in HIV testing services and male-focused interventions [13].

The observed increase in HIV testing uptake coincides with the expansion of provider-initiated testing, community-based HIV testing, voluntary medical male circumcision (VMMC), and broader HIV prevention programming implemented nationally during the past decade [25]. The slight decline in 2024 may reflect programmatic saturation, shifting testing strategies toward targeted populations, or post–COVID-19 service disruptions, as reported in other settings [27]. Nonetheless, the sustained high uptake suggests that gains achieved since 2013 have largely been maintained. Pronounced regional disparities in HIV testing uptake were evident across survey years. Western, Southern, and Lusaka provinces consistently recorded higher testing coverage, while Northern, Muchinga, and North-Western provinces lagged behind. These differences likely reflect variation in health service availability, urbanization, HIV programme intensity, population mobility, and socio-economic conditions across provinces [18,22]. Importantly, regional effects remained significant even after adjusting for individual-level characteristics in the multilevel model, underscoring the role of geographic and health-system factors in shaping access to HIV testing services.

Age emerged as a strong and consistent determinant of HIV testing uptake. Men aged 25–49 years had significantly higher odds of testing compared with younger men aged 15–24 years across all survey waves. While testing increased across all age groups between 2007 and 2018, the decline observed in 2024 was most pronounced among younger men. Similar age-related gradients have been documented in Malawi, Uganda, and multi-country DHS analyses, where younger men are less likely to perceive themselves at risk and have fewer routine interactions with health services [15,19]. These findings highlight persistent gaps in reaching adolescent and young adult men and underscore the need for youth-friendly and age-tailored HIV testing strategies.

Educational attainment consistently emerged as one of the strongest predictors of HIV testing uptake. Men with secondary or higher education had substantially higher odds of testing than those with no formal education, even after controlling for other covariates. This association likely reflects improved health literacy, greater awareness of HIV-related risks, and enhanced ability to navigate health systems among more educated men [8,21,28]. While long-term investments in education remain essential, short-term HIV programmes should prioritise simplified, culturally appropriate messaging and community outreach to men with limited formal education. Employment status and household wealth were also positively associated with HIV testing uptake. Employed and wealthier men were more likely to have tested for HIV, consistent with evidence from Burkina Faso and other African settings showing that economic stability facilitates access to health services and reduces indirect costs associated with testing [22,29,30]. These findings suggest that workplace-based testing, flexible service hours, and community outreach targeting economically vulnerable men may help address persistent inequalities in testing uptake.

Marital status was among the strongest determinants of HIV testing uptake, with married men exhibiting more than fourfold higher odds of testing compared with unmarried men. This likely reflects increased exposure to couple-based testing through antenatal care, partner encouragement, and the expansion of HIV self-testing within households [31,32]. However, qualitative evidence from Uganda and other settings indicates that traditional masculinity norms and fears of disclosure may deter some men from engaging in couple testing [23]. These findings highlight the need for male-sensitive counselling approaches that address gender norms while promoting shared responsibility for HIV prevention.

HIV knowledge demonstrated a clear dose–response relationship with HIV testing uptake, with progressively higher odds of testing observed as knowledge scores increased. This finding is consistent with regional evidence indicating that knowledge plays a central role in shaping health-seeking behaviour and engagement with HIV services [29,33]. Media exposure was similarly associated with higher testing uptake, reinforcing the importance of mass media as a channel for disseminating HIV prevention information and service availability [33]. Notably, awareness of sexually transmitted infections (STIs) was positively associated with HIV testing uptake in this study, contrary to some earlier findings. Men who were aware of STIs were more likely to have tested for HIV, potentially reflecting higher risk perception and increased interaction with sexual and reproductive health services [34]. This suggests that integrating HIV testing with STI education and services may offer an effective pathway for increasing male testing uptake.

Across all survey years, HIV testing uptake increased substantially among both urban and rural men, although persistent disparities by place of residence were evident. In 2007, HIV testing uptake was low in both settings but markedly lower among rural men compared with urban men, reflecting long-standing inequities in access to HIV services in Zambia and other sub-Saharan African countries [1,2]. Substantial improvements were observed in 2013–14 and 2018, coinciding with the national scale-up of provider-initiated testing, community-based HIV testing, and outreach services that expanded testing beyond health facilities [3,4,14]. Despite these gains, urban men consistently reported higher testing uptake than rural men across all survey years. By 2018, HIV testing uptake exceeded 80% among urban men and reached approximately 75% among rural men. In 2024, a slight decline was observed in both groups; however, urban men continued to report higher uptake than rural men (76.2% vs. 71.1%), and levels in both settings remained substantially higher than those recorded in 2007.

These temporal patterns align with the multilevel findings showing that rural residence was independently associated with lower odds of HIV testing compared with urban residence. Similar rural–urban disparities have been widely documented across sub-Saharan Africa, where men living in rural areas face structural barriers such as limited availability of health facilities, longer travel distances, transportation costs, reduced exposure to mass media campaigns, and fewer opportunities for workplace- or venue-based HIV testing [2,6,15,35]. In addition, concerns about confidentiality and fear of stigma in closely knit rural communities may further discourage men from seeking HIV testing [9,15]. Together, these findings indicate that while national HIV testing strategies have improved coverage in both urban and rural settings, geographic access and service-delivery constraints continue to disadvantage rural men. Strengthening decentralised, mobile, and male-friendly HIV testing approaches tailored to rural contexts remains essential for achieving equitable HIV testing coverage among men in Zambia.

In the fully adjusted multilevel model, community-level poverty, education, and employment were not significantly associated with HIV testing uptake, and residual clustering at the community level was minimal. This suggests that individual-level socio-demographic characteristics exert greater influence on HIV testing behavior among men in Zambia than aggregated community factors. Similar findings have been reported in recent studies from South Africa, where individual determinants outweighed community-level effects after adjustment [22,33]. Nonetheless, persistent regional differences indicate that broader structural and health-system factors remain important and warrant targeted policy attention.

### Policy Recommendation

Sustaining and accelerating progress in HIV testing among men in Zambia requires targeted, male-centered strategies aligned with the priorities of the **Zambia National HIV Strategic Framework** under the leadership of the Ministry of Health Zambia. Policy efforts should prioritize younger men and rural populations through decentralized, mobile, and venue-based HIV testing services, while expanding access to HIV self-testing and integrating testing into voluntary medical male circumcision (VMMC), workplace, and family-centered programmes. Strengthening HIV education and tailored communication for men with lower educational attainment remains critical, given the strong association between HIV knowledge and testing uptake. Region-specific implementation plans should address persistent provincial disparities to ensure equitable coverage. Collectively, these approaches will support early diagnosis, timely linkage to care, and sustained progress toward national and global HIV targets.

### Strength and Limitations

This study has several important strengths. First, it used nationally representative data from four rounds of the Zambia Demographic and Health Surveys (ZDHS) spanning 2007 to 2024, enabling robust assessment of long-term trends in HIV testing uptake among men in Zambia. The large sample size and standardized DHS methodology enhance the reliability and comparability of findings across survey years. Second, the application of survey weights and adjustment for the complex sampling design ensured nationally representative estimates and minimized bias arising from unequal selection probabilities. Third, the use of multilevel logistic regression allowed for appropriate accounting of the hierarchical structure of the data and provided more precise estimates of associations between HIV testing uptake and individual-level determinants. Finally, the inclusion of a wide range of socio-demographic and behavioural variables enabled a comprehensive examination of factors associated with HIV testing uptake among men.

Despite these strengths, several limitations should be acknowledged. The cross-sectional nature of the DHS data precludes causal inference, and the observed associations should therefore be interpreted as correlational rather than causal. HIV testing history was self-reported and may be subject to recall bias or social desirability bias, potentially leading to over-reporting of testing uptake. In addition, some relevant factors influencing HIV testing behaviour among men—such as perceived stigma, masculinity norms, health system quality, and availability of HIV self-testing were not consistently captured across all survey waves and could not be examined in this analysis. Although multilevel modelling was employed, unmeasured contextual or programmatic factors at the community or facility level may still influence HIV testing uptake. Finally, slight variations in questionnaire content and variable definitions across survey years may have introduced minor inconsistencies, although DHS harmonization procedures were applied to minimize such effects.

## Data Availability

The data underlying the results presented in this study are publicly available from the Demographic and Health Surveys (DHS) Program repository. Specifically, the Zambia Demographic and Health Survey (ZDHS) datasets for the years 2007, 2013/14, 2018, and 2024 can be accessed at https://www.dhsprogram.com following user registration and submission of a formal data request. Access to the datasets is granted upon approval by the DHS Program, and the data are provided in anonymized form to protect the confidentiality of survey respondents.

## Acknowledgement

The authors gratefully acknowledge the Zambia Statistics Agency, the Ministry of Health Zambia, and the ICF for granting access to the Zambia Demographic and Health Survey (ZDHS) datasets used in this study. We also extend our appreciation to all survey participants for their time and valuable contributions, without which this research would not have been possible. The technical and logistical support provided through the Demographic and Health Surveys (DHS) Program is sincerely acknowledged.

## Clinical Trial Number

Not applicable

## References

1. UNAIDS. Global HIV & AIDS statistics—Fact sheet. Geneva: Joint United Nations Programme on HIV/AIDS; 2024. https://www.unaids.org/en/resources/fact-sheet#:~:text=40.8%20million%20%5B37.0%20million%E2%80%9345.6,AIDS%2Drelated%20illnesses%20in%202024.

2. UNAIDS. The Global AIDS Update 2024. Geneva: Joint United Nations Programme on HIV/AIDS; 2024. https://www.unaids.org/en/resources/documents/2024/global-aids-update-2024.

3. Zambia Statistics Agency (ZamStats), Ministry of Health (MOH) Zambia, ICF. Zambia Demographic and Health Survey 2018. Lusaka, Zambia, and Rockville, Maryland, USA; 2019. https://dhsprogram.com/pubs/pdf/fr361/fr361.pdf.

4. Hlongwa M, Mashamba-Thompson T, Makhunga S, Hlongwana K. Barriers to HIV testing uptake among men in sub-Saharan Africa: a scoping review. Afr J AIDS Res. 2020;19(1):13–23. doi:10.2989/16085906.2020.1725071.

5. Madut DB, et al. Predictors of prior HIV testing and acceptance of a community-based HIV test offer among male bar patrons in northern Tanzania. PLOS Glob Public Health. 2024;4(2):e0002946.

6. Seidu AA. Using Anderson’s Model of Health Service Utilization to Assess the Use of HIV Testing Services by Sexually Active Men in Ghana. Front Public Health. 2020 Sep 15;8:512. doi: 10.3389/fpubh.2020.00512. PMID: 33042949; PMCID: PMC7522213.

7. Hensen B, et al. Frequency of HIV-testing and factors associated with multiple lifetime HIV-testing among a rural population of Zambian men. BMC Public Health. 2015;15:960. doi:10.1186/s12889-015-2259-3.

8. Adugna DG, Worku MG. HIV testing and associated factors among men (15–64 years) in Eastern Africa: a multilevel analysis using recent DHS data. BMC Public Health. 2022;22:2170. doi:10.1186/s12889-022-14588-6.

9. Ha JH, et al. Gendered relationship between HIV stigma and HIV testing among men and women in Mozambique: a cross-sectional study. BMJ Open. 2019;9:e029748.

10. Muchimba M, Zyambo C. Characteristics and sexual risk behaviour of men who never tested for HIV in Zambia. Am J Mens Health. 2021;15(6):15579883211063343. doi:10.1177/15579883211063343.

11. Akweh TY, Boakye BAJ, Adoku E, Teyko F, Tarkang EE. HIV testing uptake and associated factors among Ghanaian men: evidence from the 2022 DHS. Discov Public Health. 2025;22:310. doi:10.1186/s12982-025-00609-3.

12. Odii IO, Chipalo E. Relationship between HIV testing and HIV transmission risk behaviours among men in Cameroon. Texila Int J Public Health. 2024;12(1):71.

13. Makusha T, Mabaso M, Richter L, Desmond C, Jooste S, Simbayi L. Trends in HIV testing and associated factors among men in South Africa. Public Health. 2017;143:1–7. doi:10.1016/j.puhe.2016.10.017.

14. Sharma M, Ying R, Tarr G, Barnabas R. Systematic review and meta-analysis of community- and facility-based HIV testing to address linkage gaps in sub-Saharan Africa. Nature. 2015;528:S77–S85.

15. Nangendo J, Obai G, Apili G, Odongo S. Prevalence, associated factors and perspectives of HIV testing among men in Uganda. PLOS ONE. 2020;15(8):e0237402. 10.1371/journal.pone.0237402

16. Ng’ambi W, Chiumia IK, Chagoma N, Mfutso-Bengo J. Factors associated with uptake of HIV testing in Malawi: trend analysis of DHS data (2004–2016). J HIV AIDS Res. 2020;2(1):101.

17. Budu E, et al. What has comprehensive HIV/AIDS knowledge got to do with HIV testing among men in Kenya and Mozambique? J Biosoc Sci. 2022;54(4):558–571.

18. Lealem EB, Zeleke EG, Andargie BA, Wagnew A. Pooled prevalence, spatial variation and factors associated with HIV testing uptake in sub-Saharan Africa. PLOS ONE. 2024;19(7):e0306770.

19. Mandiwa C, Namondwe B. Uptake and correlates of HIV testing among men in Malawi. BMC Health Serv Res. 2019;19:203. doi:10.1186/s12913-019-4031-3.

20. Jude O, Nelson O, Katagwa I. Socio-economic and demographic factors associated with never having tested for HIV among sexually active men in Uganda. BMC Public Health. 2021;21:2301. doi:10.1186/s12889-021-12384-2.

21. Adeagbo O, Xulu Z, Gumede D, Naidoo K. Barriers and strategies to improve men’s uptake of HIV care services in rural KwaZulu-Natal, South Africa. J Law Soc Dev. 2024. doi:10.25159/2520-9515/14877.

22. Stephenson R, Elfstrom KM, Winter A. Community influences on married men’s uptake of HIV testing in eight African countries. AIDS Behav. 2013;17(7):2352–2366. doi:10.1007/s10461-012-0223-0.

23. Siu GE, Wight D, Seeley JA. Masculinity, social context and HIV testing: an ethnographic study of men in rural eastern Uganda. BMC Public Health. 2014;14:33. doi:10.1186/1471-2458-14-33.

24. Muchimba M, Zyambo C. Fatherhood and HIV testing among men in Zambia. Am J Mens Health. 2021;15(6):15579883211063343. doi:10.1177/15579883211063343.

25. Voluntary medical male circumcision program in Zambia. 2023. https://www.clintonhealthaccess.org/case-study/voluntary-medical-male-circumcision-program-in-zambia-expands-despite-covid-19/?utm_source=chatgpt.com.

26. JAIDS Journal of Acquired Immune Deficiency Syndromes. 2026. https://journals.lww.com/jaids/pages/default.aspx.

27. Disruption in services for HIV, viral hepatitis and sexually transmitted infections during the COVID-19 pandemic in the WHO European Region: a scoping review. 2022. https://www.who.int/europe/publications/i/item/9789289058384.

28. Wandera SO, Kwagala B, Maniragaba F. Prevalence and determinants of recent HIV testing among older persons in rural Uganda: a cross-sectional study. BMC Public Health. 2020;20(144).

29. Ijaiya MA, Anibi A, Abubakar MM, Obanubi C, Anjorin S, Uthman OA. A multilevel analysis of the determinants of HIV testing among men in sub-Saharan Africa: evidence from Demographic and Health Surveys across 10 African countries. PLOS Glob Public Health. 2024;4(5):e0003159. doi:10.1371/journal.pgph.0003159.

30. De Allegri M, Agier I, Tiendrebéogo J, Louis VR, Yé M, Mueller O, Ridde V. Factors affecting the uptake of HIV testing among men: a mixed-methods study in rural Burkina Faso. PLOS ONE. 2015;10(7):e0130216. doi:10.1371/journal.pone.0130216.

31. Sundararajan R, Ponticiello M, Nansera D, Jeremiah K, Muyindike W. Interventions to increase HIV testing uptake in global settings. Curr HIV/AIDS Rep. 2022;19(3):184–193. doi:10.1007/s11904-022-00602-4.

32. Jooste S, Mabaso M, Taylor M, North A, Shean Y, Simbayi LC. Socio-economic differences in the uptake of HIV testing and associated factors in South Africa. BMC Public Health. 2021;21(1):1591. doi:10.1186/s12889-021-11583-1.

33. Awopegba OE, Ologunowa TO, Ajayi AI. HIV testing and self-testing coverage among men and women in South Africa: an exploration of related factors. Trop Med Int Health. 2021;26(2):214–227. doi:10.1111/tmi.13514.

34. Maviso M. Prevalence and predictors of HIV testing among young men in Papua New Guinea: a cross-sectional analysis of a nationally representative sample. PLOS ONE. 2024;19(8):e0306807. doi:10.1371/journal.pone.0306807.

35. Madut DB, Achiro L, Joseph B, Mbabazi P, Kagaayi J, Ssekubugu R, et al. Predictors of prior HIV testing and acceptance of a community-based HIV test offer among male bar patrons in northern Tanzania. PLOS Glob Public Health. 2024;4(2):e0002946. doi:10.1371/journal.pgph.0002946.

